# Transcriptomic Profiling and Demographic Analysis of Appalachian Patients with Pancreatic Ductal Adenocarcinoma using the Purity Independent Subtyping of Tumors (PurIST) Classifier

**DOI:** 10.64898/2026.09.22.26363302

**Authors:** Chelsey M. Williams, Reema A. Patel, Nicole Lisek, Chandra Kakarala, Michael J. Cavnar, Joseph Kim, Prakash K. Pandalai, Emily Baiyee Toegel, Tiago Biachi de Castria, Ashish Manne, Robert J. Rounbehler, Patrick M Boland, Carlos H. F. Chan, Muneeb Rehman, Deepak Vadehra, Dae Won Kim, Michelle L Churchman, Derek B. Allison, Justin B. Miller

## Abstract

**PURPOSE:** Pancreatic ductal adenocarcinoma (PDAC) is a deadly malignancy with poor prognosis. A transcriptomic-based algorithm called Purity Independent Subtyping of Tumors (PurIST) relies on expression profiling of a defined set of genes to stratify PDAC into classical and basal-like molecular subtypes and assigns a confidence level to the subtypes as either “strong,” “likely,” or “lean.” In this study, we apply PurIST to Appalachian and non-Appalachian patient cohorts and assess demographic variables between groups that may influence treatment outcomes.

**PATIENTS AND METHODS:** This 13-site study includes 780 PDAC patients. Patients were divided into Appalachian and non-Appalachian cohorts according to Appalachian Regional Commission guidelines, and differences in PurIST classification, tobacco use, and alcohol use were analyzed.

**RESULTS:** Individuals from Appalachian regions were more likely to receive a subtype classification from PurIST with “strong” confidence as opposed to “likely” or “lean” (*P*=0.0175; odds ratio=1.66). We did not observe any significant difference between basal and classical subtype classification in Appalachia (*P*=0.1457), yet an exploratory analysis showed that individuals aged ≥85 were significantly more likely to have basal-like tumor classifications (*P*=0.005817; odds ratio=3.79). Appalachian patients were more likely to have a history of smoking (59.6% vs 49.9%; *P*=0.01756) and less likely to report alcohol consumption (50.5% vs 65.6%; *P*=0.000229).

**CONCLUSION:** We conducted the largest comparison of the PurIST algorithm in Appalachia to date and found PurIST is more likely to have higher confidence in molecular subtype classifications for Appalachian PDAC tumors compared to non-Appalachian tumors. If PurIST is to be considered as a prognostic or decision-making tool for PDAC patients, we must account for variability in different populations and model the likelihood that tumors will have a strong molecular signature.

## Background

Pancreatic ductal adenocarcinoma (PDAC) has a 5-year survival of ~13% in the United States and remains a leading cause of cancer mortality, resulting in ~50,000 deaths per year [1]. Current frontline chemotherapy regimens include either modified 5-fluorouracil, oxaliplatin, leucovorin, and irinotecan (mFOLFIRINOX) or gemcitabine with albumin-bound paclitaxel [2]. Inconsistent treatment outcomes have driven efforts to develop biologically guided strategies for prognosis and chemotherapy selection, which rely on accurate tumor subtype classification. The Purity Independent Subtyping of Tumors (PurIST) classifier, a single-sample classifier for pancreatic cancer, relies on expression profiling of a defined set of genes to stratify PDAC tumor samples into classical and basal-like molecular subtypes [3]. In retrospective studies utilizing the PurIST classifier, patients with basal-like tumors had worse outcomes, regardless of which first-line chemotherapy was received [4, 5]. Ongoing prospective clinical trials are utilizing PurIST to guide clinical chemotherapy decisions and prognosis [6, 7]. While these studies have been conducted in large-volume retrospective cohorts, the applicability of PurIST to real-world patient cohorts in regionally defined areas such as Appalachia has not yet been established.

The Appalachian region stretches across 13 states, extending from the Mississippi Delta to southern New York [8]. Appalachian patients are often localized to rural areas, and many people live two or more hours away from tertiary care facilities and/or National Cancer Institute-designated Comprehensive Cancer Centers [9]. Although there has been a notable reduction in all-cause and cancer mortality nationwide, the decline of cancer mortality in Appalachia has occurred at a slower rate [10]. High cancer burden in the Appalachian population has been linked to health behaviors, social determinants of health, and diminished access to tertiary care facilities [10, 11]. Additional contributing factors include screening rate disparities, high obesity rates, increased tobacco use, unfavorable economic conditions, environmental exposures, and regional lack of primary care physicians [10-14]. It is unclear if population-specific molecular and transcriptomic risk factors drive prognosis and treatment response in Appalachia. Therefore, this work assesses the PurIST algorithm in Appalachia to determine if population-specific biases should be considered to ensure that the standard of care and patient outcomes are comparable in this region compared to the rest of the United States. Here, we outline differences in cancer risk factors and molecular subtypes observed between Appalachian and non-Appalachian populations.

## Methods

### Cohort Design

#### ORIEN Avatar Project

The Oncology Research Information Exchange Network (ORIEN) is an alliance of United States cancer centers established in 2014. All ORIEN alliance members utilize a standard Total Cancer Care ® (TCC) protocol or biospecimen collection protocol that share common core elements with the TCC protocol. TCC-consented patients with biospecimens available and who meet eligibility criteria may be included into the ORIEN Avatar Project, which includes research use only grade RNA sequencing and collection of deep longitudinal clinical data with lifetime follow up. 780 ORIEN Avatar patients diagnosed with PDAC and consented to the TCC protocol from the participating members of ORIEN were included in this study.

### Data Availability

The data used in this research was generated through private funding by Aster Insights (www.asterinsights.com) in collaboration with the ORIEN (www.oriencancer.org). Requests for access to the data used in this study can be submitted to the corresponding author and.

This 13-site study includes 780 PDAC patients via the TCC protocol. This protocol is a longitudinal study which collects tissue and blood biospecimens as well as clinical data via ORIEN with Aster Insights **(Figure 1; Table 1)**.

**Table 1.** Participating ORIEN institutions, separated by Appalachian and non-Appalachian regions.

| Participating ORIEN Institutions |  |
| --- | --- |
| Appalachian | Non-Appalachian |
| • Murtha Cancer Center Program<br>• Roswell Park Comprehensive Cancer Center<br>• The Ohio State University Comprehensive Cancer Center – Arthur G. James Cancer Hospital and Richard J. Solove Research Institute<br>• University of Kentucky Markey Cancer Center<br>• University of Virginia | • Indiana University Melvin and Bren Simon Comprehensive Cancer Center<br>• Moffitt Cancer Center and Research Institute<br>• Oklahoma University Stephenson Cancer Center<br>• Rutgers Cancer Institute of New Jersey<br>• University of Colorado Cancer Center<br>• University of Iowa Holden Comprehensive Cancer Center<br>• University of Kansas Cancer Center<br>• USC Norris Comprehensive Cancer Center |

**Figure 1:**
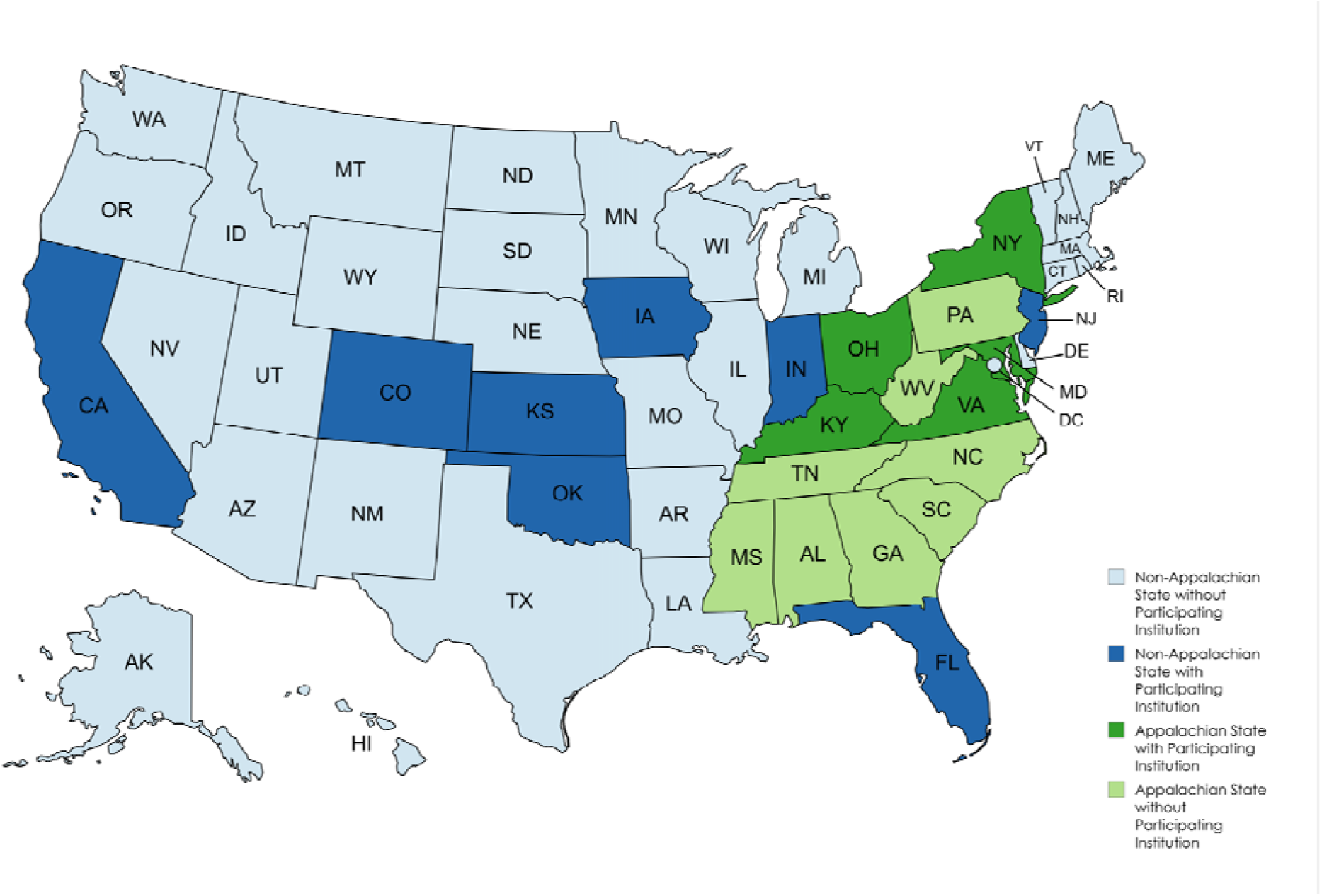
Map of the United States denoting Appalachian and non-Appalachian states according to Appalachian Regional Commission guidelines. Appalachian states with participating ORIEN institutions are dark green. Non-Appalachian states with participating ORIEN institutions are dark blue.

Although the ORIEN network includes data from patients diagnosed with cancer at any age, this study focused exclusively on adults (age ≥ 18), who were capable of signing informed consent and research authorization forms in either English or Spanish. Enrolled patients also must have received clinical care at a participating ORIEN institution and have been diagnosed with PDAC. No other inclusion criteria were used (**Table 2**).

**Table 2.** Patient inclusion criteria.

| Category | Inclusion Criteria |
| --- | --- |
| Age | Patients of any age over 18 |
| Cancer Diagnosis | Pancreatic ductal adenocarcinoma |
| Consent | Ability to understand and sign informed consent |
| Performance Status | No restrictions on Eastern Cooperative Oncology Group (ECOG) performance status |
| Germline Alterations | No restrictions based on germline mutation status |
| Disease Stage | No restrictions on cancer stage |
| Tumor Markers | No restrictions based on tumor marker status |
| Line of Therapy | No restrictions on prior, current, or line of systemic therapy |
| Patient Location | Clinically evaluated at participating ORIEN institution |

Patients with PDAC histology were divided into two cohorts, Appalachian and non-Appalachian, following guidelines from the Appalachian Regional Commission, which defined Appalachia as a 206,000-square-mile region spanning 13 states[8] **(Figure 2)**.

**Figure 2:**
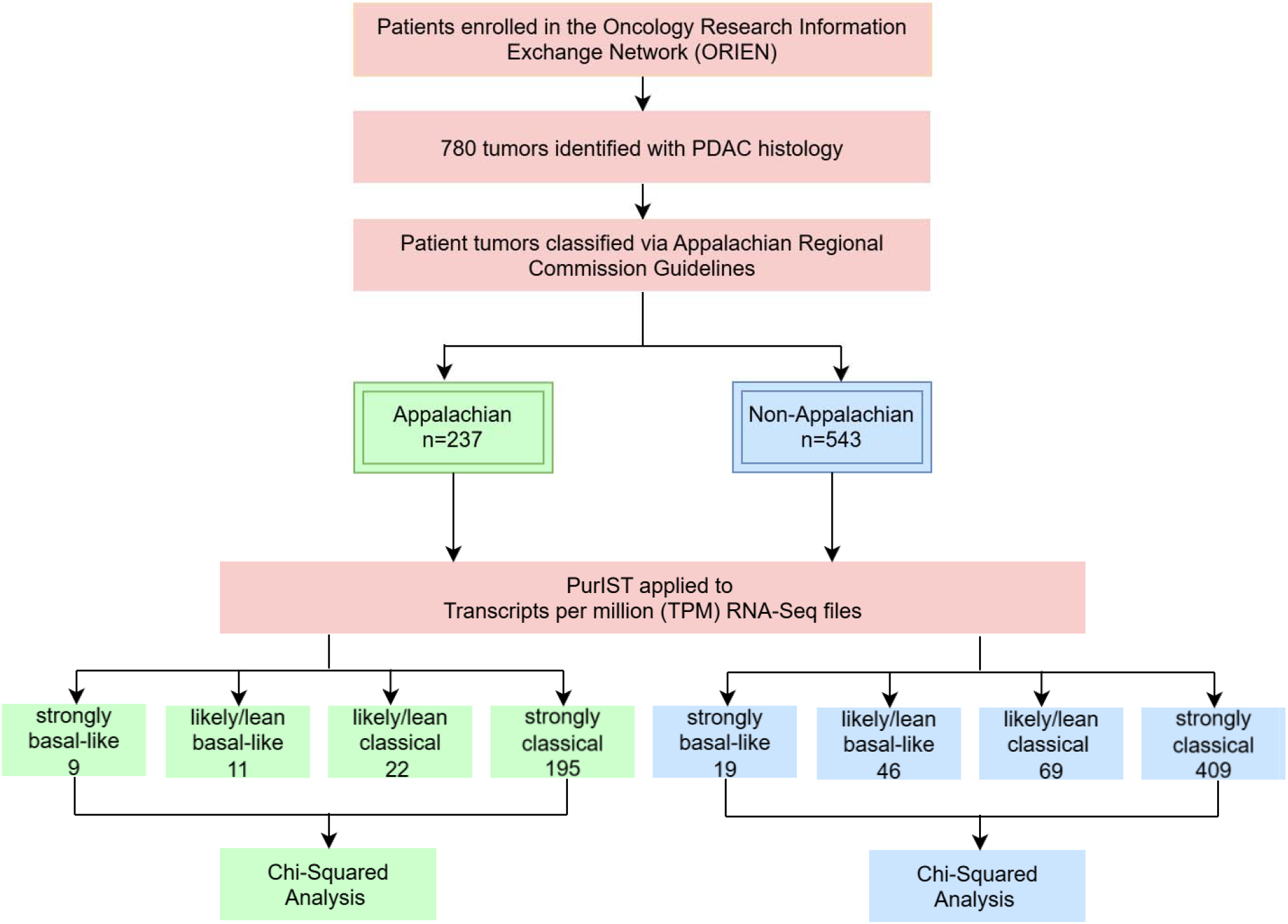
CONSORT Diagram. PurIST, purity independent subtyping of tumors. Samples that PurIST classified with confidence levels of “lean” and “likely” were combined due to the limited number of samples classified as “lean.”

Since previous studies have linked smoking and heavy drinking to differences in PDAC incidence rates [15], we used a chi-square analysis comparing differences in the self-reported incidence of tobacco and alcohol use from individuals within our cohort that reside in Appalachian compared with non-Appalachian regions.

Although there were 814 sample IDs (SLID)s, 34 individuals were sequenced multiple times. To account for sample inflation, we matched the duplicate ORIEN Avatar IDs to the SLIDs and ensured that only the first entry was included in downstream analyses.

### Running the PurIST algorithm

The PurIST algorithm includes a pre-defined list of 16 genes: *ANXA10, BCAR3, C16orf74, CLDN18, CLRN3, DDC, GATA6, GPR87, ITGA3, KRT5, KRT6A, LGALS4, PTGES, REG4, S100A2, SLC40A1*. Quality control (QC) metrics for the RNA-seq data were previously computed by Aster Insights, and **Table 3** shows the QC thresholds that were set.

**Table 3.** Quality control thresholds for RNA-seq data.

| QC Metric | PASS Threshold | FLAG threshold | FAIL threshold |
| --- | --- | --- | --- |
| Number of reads (M) passing filters | > 50 | 20 to 50 | < 20 |
| Exonic Rate | > 0.6 | 0.4 to 0.6 | < 0.4 |
| Unique reads (M) | 10 | 1 to 10 | < 1 |
| Duplication rate of mapped reads | < 0.9 | 0.9 | N/A |
| Number of genes detected | 12,500 | 5,000 to 12,500 | < 5,000 |

None of the samples failed any QC metric; however, 59 samples had a FLAG for at least one metric. The QC thresholds were set by the ORIEN network and indicate optimal sequencing metrics. A FLAG means that the sequencing had borderline QC metrics yet still passed the low QC threshold. While we recognize that the likelihood of unintended sequencing issues increases when QC metrics are in the FLAG threshold as opposed to the PASS threshold, we also wanted to evaluate a real-world scenario in which PurIST was applied to research-grade sequencing data. Therefore, we assessed if the samples with sequencing FLAGs were more likely to show outlier expression values than samples without FLAGs, which might indicate a systematic issue that could limit the PurIST algorithm from returning accurate results. We performed an outlier assessment for each of the 16 genes required by the PurIST algorithm using the transcripts per million (TPM) values. We found that 125 out of 814 total samples (including individuals who were sequenced multiple times since some, but not all, of those individuals had a FLAG for one sample and PASS for another) had outlier gene expression for at least one gene. Of those 125 samples, 12 / 125 (9.6%) had a FLAG in the QC metric column, whereas 47 / 689 (6.82%) of the samples without any outlier gene expression had a FLAG in the QC metric column. There was no significant difference between those two groups (*P*=0.2703), indicating that samples with a FLAG did not create a systematic bias in sequencing that differed from the samples with a PASS for the QC metrics.

PurIST was run using the TPM values to classify tumors on a continuum that includes strong classical, likely classical, lean classical, lean basal-like, likely basal-like, and strong basal-like. A chi-square test was used to determine if the distribution of classical and basal-like tumors observed among patients from Appalachia differed from non-Appalachian patients, if molecular subtypes differed in individuals age ≥85, and if the PurIST score confidence differed by geographic location. Odds ratios were calculated from the chi-square contingency table by first calculating the odds of each variable for each population and then calculating the ratio of those odds. For smoking and alcohol consumption, individuals with unknown values were removed from the chi-squared analyses but still reported in the tables. Additionally, the “Current” and “Former/Ever” categories were combined for the chi-squared tests yet separated for the tables.

## Results

Of the 780 patients included in the study population and assessed using PurIST, 237 were from Appalachia and 543 were from outside of the Appalachian region. While the Appalachian cohort trended toward increased classical subtype compared to the non-Appalachian cohort, those differences were not significant (*P*=0.1454). A total of 695 (89.23%) were classified as classical (604 strong classical, including 195 Appalachian and 409 non-Appalachian) and 85 (10.90%) were classified as basal-like (28 strong basal-like, including 9 Appalachian and 19 non-Appalachian) (**Table 4**).

**Table 4.**
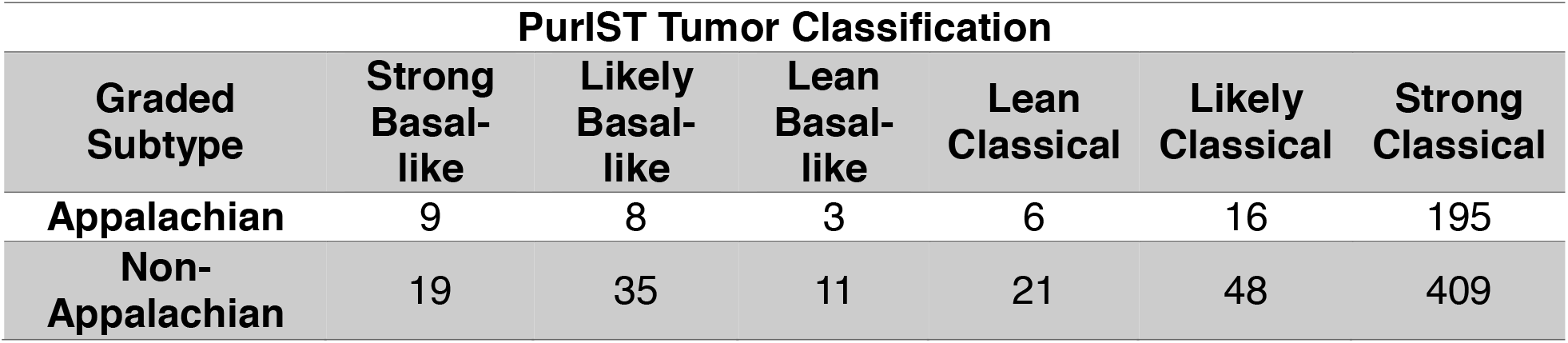
Number of tumor subtypes by region.

A chi-square test showed that individuals in Appalachia were significantly more frequently classified with a “strong” confidence in the reported molecular subtype (either classical or basal-like) compared to individuals from non-Appalachian regions; conversely, tumors of individuals from non-Appalachian regions were significantly more frequently classified with lower confidence “likely” or “lean” subtype classifications (*P*= 0.0175; odds ratio=1.66). In total, PurIST classifications with “strong” confidence designations were made for 204 out of 237 (86.1%) Appalachian and 428 out of 543 (78.8%) of non-Appalachian individuals. This study could not assess the clinical relevance of the differences between “strong” and “likely/lean” subtype classifications due to limited longitudinal data regarding clinical outcomes.

Additionally, Appalachian patients were significantly more likely to report a history of smoking (59.6% vs 49.9%; *P*=0.01756; **Table 5**) and less likely to report a history of alcohol consumption (50.5% vs 65.6%; *P*=0.000229; **Table 6**).

**Table 5.** Patient smoking status by region. Appalachian patients were significantly more likely to report a history of smoking (59.6% vs 49.9%; *P*=0.01756).

| Smoking Status |  |  |  |  |  |
| --- | --- | --- | --- | --- | --- |
|  | Current | Former/Ever | Never | Unknown/Not Applicable | Unknown/Not Reported |
| **Appalachian** | 36 | 91 | 86 | 23 | 1 |
| **Non-Appalachian** | 62 | 183 | 246 | 32 | 20 |

**Table 6.** Patient reported alcohol use by region. Appalachian patients were less likely to report alcohol consumption (50.5% vs 65.6%; *P*=0.000229).

| Alcohol Use |  |  |  |  |  |
| --- | --- | --- | --- | --- | --- |
|  | Current | Former/Ever | Never | Unknown/Not Applicable | Unknown/Not Reported |
| **Appalachian** | 71 | 30 | 99 | 23 | 14 |
| **Non-Appalachian** | 251 | 70 | 168 | 32 | 22 |

Significantly more basal-like tumors were identified in individuals age ≥85, regardless of region (**Table 7**; *P*=0.005817; odds ratio=3.79). Individuals in Appalachia had a non-significant difference in the age of last contact compared to non-Appalachian regions (Appalachian age at last contact: 66.1925 ± 12.353; non-Appalachian age at last contact: 68.0549 ± 12.0193; *P*=0.0782).

**Table 7.** Individuals age ≥85 are more likely to have a basal-like PDAC molecular tumor subtype than a classical subtype (*P*=0.005817; odds ratio=3.79).

| PDAC Tumor Subtypes by Age at Last Contact |  |  |
| --- | --- | --- |
| Age at Last Contact | Basal-Like Tumors | Classical Tumors |
| < 85 | 74 | 661 |
| ≥ 85 | 11 | 34 |

## Discussion

Our analysis includes the largest real-world evaluation of the PurIST classifier to date, and the only systematic analysis of PurIST in the Appalachian region. Appalachia is a historically understudied region with distinct socioeconomic, demographic, and environmental challenges which may influence tumor biology and treatment response [16]. Patients in Appalachia often experience substantially limited specialty cancer care, often due to geographic disparities [17, 18]. Prior PurIST evaluations have not accounted for the possibility of regional differences, and our findings begin to address this by accounting for demographic variability. Other historically underrepresented racial, ethnic, and geographically defined populations may also benefit from further PurIST analysis to assess diagnostic outcomes in each population.

When we applied the PurIST classifier to Appalachian and non-Appalachian cohorts, we found that a larger proportion of Appalachian tumors were able to be successfully classified into either strongly basal-like or strongly classical compared to non-Appalachian tumors. Significantly fewer patients were likely to be classified into lean or likely categories. One hypothesis is that this could reflect stronger subtype-defining expression patterns or greater transcriptional polarization in Appalachia, although this interpretation requires additional validation in larger cohorts. Since all samples passed QC, it is unlikely that these results can be explained by sequencing errors.

An exploratory analysis of older patients aged ≥85 showed that they were more likely to have PDAC tumors classified as basal-like compared to classical. Although this finding contrasts with prior large molecular profiling studies [4, 5] that did not identify associations of age with subtype, this raises the possibility that older patients may exhibit basal-like transcriptional characteristics.

Additionally, although known cancer risk factors such as smoking and alcohol use are more prevalent in Appalachian regions [11, 12], Appalachian PDAC patients in the ORIEN network were less likely to report alcohol use than non-Appalachian patients. While Appalachian patients were more likely to report a history of smoking than non-Appalachian patients, the difference in reporting alcohol consumption is unclear. If true, the higher PDAC incidence rate in Appalachia may be associated with additional variables beyond that of alcohol use.

Though our study included several participating institutions from the Appalachian states of Ohio, Kentucky, Virginia, New York, and Maryland, there were no participating institutions from Pennsylvania, West Virginia, Tennessee, North Carolina, South Carolina, Mississippi, Alabama, and Georgia. This is an inherent limitation of our study that cannot be overcome through the ORIEN network. Future clinical integration and data sharing policies should be explored to include those additional states, with particular attention to West Virginia, as West Virginia is the only US state in which all counties are considered Appalachian.

When evaluating our entire cohort, we found a trend towards increased expression of the classical subtype across all populations. This finding is consistent with prior published retrospective analyses [4, 5]. Unlike these studies, we did not filter our cohort by ECOG status. Prior and ongoing large-scale PurIST analyses, along with the PASS-01 trial and retrospective analysis of the COMPASS trial cohort, restrict their analyses to patients with ECOG of 0-1. Historically, individuals with ECOG 0-1 are considered candidates for intensive cytotoxic chemotherapy and are more likely to be eligible for clinical trials [2]. Restricting by ECOG status may not accurately reflect which patients could be offered PurIST as a test in the medical oncology clinical setting. Furthermore, ECOG status was not available for a proportion of our patients and thus excluding patients on the basis of ECOG from this study would have limited the statistical power of our analyses. Since PurIST is currently available to order as a test by medical oncologists in clinical practice without requiring ECOG status, we felt that our inclusion criteria most accurately reflected the real-world PurIST use case.

Our inclusion criteria were specifically designed to test population-specific differences in PurIST molecular subtyping using real-world data, which does not always include germline sequencing. Therefore, unlike previous studies [4, 5] which excluded individuals with germline alterations in *BRCA1, BRCA2, PALB2*, and other DNA damage repair genes, our study did not exclude those individuals. By casting a wider net using more inclusive criteria, our study captures clinical variability observed in routine management of patients with PDAC and is more representative of a real-world population of patients who could be offered PurIST as a clinical test.

We recognize that a statistical difference in PurIST classifications with strong confidence may not result in a clinically meaningful difference in treatment compared to individuals with a lean/likely classification. Additionally, we recognize that the ORIEN dataset has a limited number of PDAC patients from Appalachia, which may result in either false positive or false negative findings based on potential recruitment biases. However, this study presents the largest comparison of PurIST molecular subtyping in the Appalachian region to date and provides the basis for future work collecting additional transcriptomic data from both Appalachian and non-Appalachian PDAC patients to ensure that the PurIST algorithm provides equitable results to all populations.

Recent therapeutic advances in PDAC, including improved overall and progression-free survival with daraxonrasib in patients with KRAS-mutant metastatic tumors [19], heighten the need for accurate molecular stratification to guide treatment selection. Although our study was not designed to evaluate treatment outcomes, the observed regional differences in PurIST classification confidence suggest that population-specific variation may influence how molecular subtyping performs across patient groups. Future studies should assess whether treatment response differs between tumors assigned “strong” versus “likely/lean” subtype classifications and whether current molecular subtyping approaches adequately stratify PDAC risk and therapeutic benefit in Appalachian and other understudied populations.

## Conclusion

We conducted the largest comparison of the PurIST algorithm in Appalachia to date. Although our analyses still included a relatively small sample size from Appalachia, we found that Appalachian tumors were significantly more likely than non-Appalachian tumors to receive a strong-confidence PurIST classification, whether classical or basal-like. We also found that older patients aged ≥85 were significantly more likely to exhibit the basal-like tumor subtype, although future work should validate this finding given the limited sample size in our cohort. Across all cohorts, there was a consistent trend toward increased expression of the classical subtype, which aligns with findings from prior retrospective analyses. Importantly, our dataset reflects the diversity and clinical variability seen in real-world patient populations, rather than narrowly defined clinical trial cohorts. These results underscore the importance of considering population-specific factors when implementing PurIST as a prognostic or treatment selection tool for PDAC patients. Future research should continue to address regional and demographic variability to ensure effective clinical decision-making.

